# Impaired glucose metabolism in patients with diabetes, prediabetes and obesity is associated with severe Covid-19

**DOI:** 10.1101/2020.06.04.20122507

**Authors:** Stephen M. Smith, Avinash Boppana, Julie A. Traupman, Enrique Unson, Daniel A. Maddock, Kathy Chao, David P. Dobesh, Ruth I. Connor

## Abstract

**Background:** Identification of risk factors of severe Covid-19 is critical for improving therapies and understanding SARS-CoV-2 pathogenesis.

**Methods:** We analyzed 184 patients hospitalized for Covid-19 in Livingston, New Jersey for clinical characteristics associated with severe disease.

**Results:** The majority of Covid-19 patients had diabetes mellitus (DM) (62.0%), Pre-DM (23.9%) with elevated FBG, or a BMI > 30 with normal HbA1C (4.3%). SARS-CoV-2 infection was associated with new and persistent hyperglycemia in 29 patients, including several with normal HbA1C levels. Forty-four patients required intubation, which occurred significantly more often in patients with DM as compared to non-diabetics.

**Conclusions:** Severe Covid-19 occurs in the presence of impaired glucose metabolism in patients with SARS-CoV-2 infection. The association of dysregulated glucose metabolism and severe Covid-19 suggests a previously unrecognized manifestation of primary SARS-CoV-2 infection. Exploration of pathways by which SARS-CoV-2 impacts glucose metabolism is critical for understanding disease pathogenesis and developing therapies.

## INTRODUCTION

SARS-CoV-2, the causative agent of coronavirus disease 2019 (Covid-19) was first detected in late December 2019 in Wuhan province in China and rapidly spread across the globe.(1) To date over 4.7 million cases of Covid-19 have been reported worldwide with approximately 1.5 million cases in the United States and 90,340 deaths.(2,3)

Early reports from China and later Italy examined risk factors for severe Covid-19 and identified advanced age as a major indicator for increased mortality.(4,5) A recent study of over 4,000 patients with confirmed Covid-19 in the United States found older age (>65 years), obesity (BMI > 40), chronic kidney disease and a history of heart failure were most associated with hospitalization, while critical illness was linked to low oxygen saturation (< 88%) at admission, first d-dimer (>2500), first ferritin (>2500) and first C-reactive protein (>200) indicating hypoxia and inflammation in patients with clinically progressive disease.(6)

A number of studies have identified an increased risk of severe disease in Covid-19 patients with underlying health conditions. Data compiled by the COVID-19 Associated Hospitalization Surveillance Network (COVID-NET) identified hypertension (49.7%), obesity (48.3%), chronic lung disease (34.6%), diabetes mellitus (DM) (28.3%) and cardiovascular disease (27.8%) as the most commonly found co-morbidities among hospitalized Covid-19 patients in the United States.(3) A recent study of Lopinavir-Ritonavir in adults hospitalized with severe Covid-19 found 13% of patients had DM, reinforcing early observations that diabetes is a risk factor for more severe disease.(7) This is supported by data from a study of 24 patients hospitalized for Covid-19 in nine Seattle-area hospitals in which 58% of critically ill patients had DM and an average BMI of 33.(8) Interestingly, in the 2003 SARS-CoV outbreak in China, hyperglycemia and DM were also noted as risk factors for mortality and morbidity.(9) These observations and several in-depth reviews(10–12) have raised concerns that diabetics with elevated fasting blood glucose are at increased risk of developing severe Covid-19.

We report here our experience of 184 patients admitted for Covid-19 to a teaching hospital in Livingston, New Jersey within the epicenter of the SARS-Cov-2 pandemic in the United States. Extending early observations, we find the vast majority of our Covid-19 patients are diabetic, prediabetic or obese. Moreover, we identify Covid-19 patients with PreDM and others with normal HbA1C levels who developed new onset DM, similar in presentation to Type 1 DM, coincident with recent acquisition of SARS-CoV-2 infection. Our data establish that impaired glucose metabolism, due to either DM or obesity, is significantly associated with severe Covid-19 in this high-risk population.

## METHODS

### Study Population

Patients with Covid-19 were referred to our practice by the Emergency Medicine department or admitting physician at a large suburban hospital (Saint Barnabas Medical Center, Livingston, New Jersey). Consecutive admission of 184 Covid-19 patients occurred over a period of seven weeks (March 16, 2020 to May 2, 2020) and all patients received care through our practice. A diagnosis of Covid-19 was made on the basis of a confirmed positive laboratory test for SARS-CoV-2 in 177 patients. The remaining patients were diagnosed based on clinical presentation including new onset hypoxia, increased LDH, increased D-dimer, increased ferritin and elevated blood glucose. All patients were included in the analysis for this study. Ethical approval for the study was granted by the Institutional Review Board of St. Barnabas Medical Center.

### Diabetes Status

A high percentage of patients testing positive for SARS-CoV-2 and referred to our practice were already known diabetics and receiving treatment for DM at the time of admission. We used the ADA definitions to diagnose DM, New Onset DM and PreDM.(13) A new diagnosis of DM was made in patients previously unaware of their condition based on an HbA1C > 6.4%. New onset DM was defined by persistently elevated fasting blood glucose (FBG) > 125 mg/dL and requiring insulin therapy. Prediabetes (PreDM) was defined by an HbA1C of 5.7 – 6.4%. Non-diabetic patients were defined as having an HbA1C < 5.7% and FBG ≤ 125 mg/dL. Fever was defined as Tmax ≥100°F during the first 6 hrs after admission. Hypoxia was defined as room oxygen saturation < 94%.

### Outcomes

The primary indicator of severe Covid-19 was intubation. The need for intubation was determined on the basis of clinical presentation in patients receiving full care throughout their hospitalization. Death during hospitalization included patients put on comfort care at any time during or after admission. Comfort care measures were determined by the primary attending physician and included but were not limited to morphine drips or intensive care without further escalation of care.

### Statistical Analyses

A one-sample proportion Z-test was used to determine the prevalence of DM, PreDM, and NonDM in Covid-19 patients as compared to the US population. The sample size used for this analysis was 184 with at least 10 patients in each DM status. One-sided hypothesis tests were used to determine if the proportions of Covid-19 patients with DM and PreDM were larger than the U.S. population proportions, and if the proportion of NonDM patients was smaller than the U.S. population proportion. A chi-squared test was used to determine significance between intubation and diabetes status within each patient group. 95% confidence intervals were calculated using standard errors. Statistical significance was defined as a P-value < 0.05. All statistical analyses were performed using R version 3.4.4.

## RESULTS

### Demographic and clinical characteristics of the patients

During a seven-week period, 184 patients were admitted to the hospital for Covid-19 and referred to our practice. The average age of study patients was 64.4 years (range: 21–100 yrs.) with 86 (46.7%) females and 98 (53.3%) males (Table 1). The racial and ethnic composition of the study population was black (53.8%), white (25.5%), Latino (6.5%) and Asian (6.0%). Clinical presentation at the time of admission included hypoxia (83.7%) and fever (62.5%) (Table 1). Hypoxia and fever occurred together often (48.9%); only a small percentage (7.6%) of patients presented without fever or hypoxia. The most common preexisting conditions included hypertension (60.3%), hyperlipidemia (33.7%), dementia (13.0%),chronic kidney disease (13.0%) coronary artery disease (12.0%), and congestive heart failure (10.9%) (Table 1).

**Table 1.** Demographic and clinical characteristics of Covid-19 patients.

| **Characteristic** | **Patients** |
| --- | --- |
| **Age** | N=184 |
| Ave- Years | 64.4 (21-100) |
| Age - No. (%) |  |
| ≤60 | 75 (40.8) |
| >60 | 109 (59.2) |
| **Gender - No. (%)** |  |
| Female | 86 (46.7) |
| Male | 98 (53.3) |
| **Race and Ethnicity-No. (%)** |  |
| Black | 99 (53.8) |
| White | 47 (25.5) |
| Latino | 12 (6.5) |
| Asian | 11 (6.0) |
| Other | 15 (8.2) |
| **Presentation Values** |  |
| BMI - Ave (Range) | 29.8 (17.5-61.4) |
| HbA1C - Ave % (Range) | 7.4 ( 4.1-14.7) |
| Glucose - Ave mg/dL (Range) | 179.9 (11-1129) |
| T ≥100°F - No. (%) | 115 (62.5) |
| Hypoxia on room air - No. (%) | 154 (83.7) |
| **Preexisting Conditions** | **<u>No. (%)</u>** |
| Hypertension | 111 (60.3%) |
| Hyperlipidemia | 62 (33.7%) |
| Dementia | 24 (13.0%) |
| Chronic Kidney Disease | 24 (13.0%) |
| Coronary Artery Disease | 22 (12.0%) |
| Congestive Heart Failure | 20 (10.9%) |
| Asthma | 18 (9.8%) |
| Atrial Fibrillation | 14 (7.6%) |
| Cancer | 17 (9.2%) |
| COPD | 12 (6.5%) |
| Cerebrovascular Accident | 10 (5.4%) |
| Transplant, Renal | 5 (2.7%) |

### Increased prevalence of diabetes, pre-diabetes and obesity

The majority of Covid-19 patients had DM (62.0%), PreDM (23.9%) or BMI > 30 with normal HbA1C (4.3%). The prevalence of DM was 4.7-fold higher in this patient group as compared to the general US population, while the prevalence of PreDM was 1.3-fold higher.(14) A significant number of patients were clinically obese. The mean BMI of the study patients was 29.8 (17.5–61.4), including 20 patients with BMIs > 40. HbA1C levels measured at admission in 171 patients also showed significant elevation with 64 patients (37.4%) having values between 5.7–6.4% and 82 (48.0%) having values ≥ 6.5%.

### Age relationship to BMI, HbA1C and initial blood glucose level

To determine whether patient age was associated with differences in clinical presentation, data on BMI, HbA1C and initial FBG were stratified by age at admission. The rates of DM and PreDM were similar in patients ≤60 yrs as compared to those > 60 yrs (Table 2) as were mean initial FBG levels (200.5 vs 165.4 mg/dL). However, patients ≤60 years of age were significantly more likely to be clinically obese. As compared to patients > 60 yrs, the frequency of obesity and the mean BMI in those ≤60 yrs were significantly higher (26.6% vs 65.3% and 27.2 vs 33.4, respectively; *p < 0.0001*) (Table 3). Patients ≤ 60 yrs were also significantly more likely to be severely obsese with a BMI>40 (20.0% vs 3.7%, p = 0.0013). Similarly, patients ≤ 60 yrs had a significantly higher mean HbA1C level than older patients (8.0 vs 6.9%; *p = 0.003*) suggesting more pronounced metabolic dysregulation in younger patients. Taken together, these data indicate that younger patients may be more likely to present with abnormalities in glucose metabolism due to obesity, which may put them at increased risk of developing severe Covid-19. These findings are consistent with a recent report of 265 Covid-19 patients demonstrating a significant inverse correlation of age and BMI in which younger patients hospitalized for Covid-19 were more often obese.(15)

**Table 2:** Diabetes status, obesity rate and HbA1C levels by age.

|  | Patients ≤60 yrs<br>(N=75) | Patients >60 yrs<br>(N=109) | *p-value* |
| --- | --- | --- | --- |
| DM (%)<sup>1</sup> | 64.0 | 60.6 | 0.38 |
| PreDM (%)<sup>1</sup> | 24.0 | 23.9 | 0.50 |
| NonDM (%)<sup>1</sup> | 12.0 | 15.6 | 0.32 |
| Mean FBG (mg/dL)<sup>2</sup> | 200.5 | 165.4 | 0.10 |
| Mean BMI<sup>3</sup> | 33.4 | 27.2 | <0.0001 |
| BMI>30 (%) | 65.3 | 26.6 | <0.0001 |
| BMI>40 (%) | 20.0 | 3.7 | 0.0004 |
| HbA1C (%) | 8.0 | 6.9 | 0.003 |

<sup>1</sup>Percent of patients with DM, Diabetes Mellitus; PreDM, Pre-Diabetes; NonDM, Non-Diabetic
<sup>2</sup>FBG, Fasting Blood Glucose
<sup>3</sup>BMI, Body Mass Index

**Table 3:** Hyperglycemia and obesity in Covid-19 patients requiring intubation.

|  | Intubated | Not<br>Intubated | *p-value*<br>(95% C.I.) |
| --- | --- | --- | --- |
| N (%) | 44 (23.9) | 144 (76.1) |  |
| Mean BMI<sup>1</sup> | 32.2 | 29.3 | 0.030<br>(0.3-5.8) |
| Mean HbA1C (%) | 8.0 | 7.2 | 0.034<br>(0.07-1.67) |
| Mean FBG (mg/dL)<sup>2</sup> | 238.0 | 163.7 | 0.013<br>(9.02-135.9) |

<sup>1</sup>BMI, Body Mass Index
<sup>2</sup>FBG, Fasting Blood Glucose

### Association of obesity and uncontrolled glycemia with intubation

Intubation was evaluated as an indicator of Covid-19 progression and severity in hospitalized patients. To determine whether higher rates of intubation were associated with uncontrolled glycemia, data on BMI, HbA1C and FBS were evaluated for intubated patients and compared to their non-intubated counterparts. Among 184 hospitalized patients receiving full care for Covid-19, 44 (23.9%) required intubation. The mean BMI of patients requiring intubation was significantly higher than that of non-intubated patients (32.3 vs 29.3; *p = 0.030*; 95% C.I. = 0.3–5.8). More strikingly, patients with a BMI > 40 were intubated at a significantly higher rate than patients with BMIs < 25 (47.4 vs 15.6%; *p = 0.0078*).

HbA1C levels were available for 41 intubated patients and revealed only four (9.8%) had normal values. Of these, three were known to be diabetic and receiving treatment for DM. In total, 40 of 41 (97.6%) intubated patients had either elevated HbA1C or were receiving therapy for DM. As compared to patients not requiring intubation, the mean HbA1C of intubated patients was significantly higher (8.0 vs 7.2%; *p = 0.034*; C.I. = 0.07–1.67). Accordingly, the rate of intubation among patients with poorly-controlled DM (HbA1C ≥7.5%) was significantly higher than that of patients with HbA1C < 7.5% (31.5 vs 17.8%; *p = 0.045*). The mean FBG at admission for intubated patients was also significantly increased when compared to that of patients not requiring intubation (238.0 vs 163.7 mg/dL; *p = 0.013*; C.I. = 9.02–135.9) suggesting that uncontrolled glycemia, due to obesity or DM, is a significant risk factor for severe Covid-19.

### Intubation rate increases in Non-DM, PreDM and DM patients

To determine whether Covid-19 severity was associated with diabetes status, patients requiring intubation were stratified as Non-DM, PreDM or DM. Determination of diabetic status was made on the basis of clinical presentation, HbA1C values and FBG levels for all patients. The majority (35 of 44; 79.5%) of intubated patients had DM, including seven newly diagnosed and five with new onset DM. Another seven (15.9%) were PreDM with high FBG levels. Only one patient requiring intubation was non-DM with normal HbA1C and FBG levels at admission, but was clinically obese with a BMI > 30.

Within the entire Covid-19 patient cohort, diabetes status was also associated with increasing rates of intubation. Among 25 patients with no diabetes and normal HbA1C levels, only one required intubation. Of patients with preDM, 18.5% (10 of 54) were intubated, while 28.8% (30 of 104) of patients with DM required intubation. Comparison of intubation rates demonstrated a significant difference between non-DM patients and those with DM (28%; *p* = *0.0094*: C.I. = 7.4–34.7%). These findings again suggest that changes in glycemia occurring with the onset and progression of diabetes are associated with a corresponding increase in the likelihood of severe Covid-19 requiring intubation.

Twenty-four patients died without intubation. The average age of these patients was 80.5 yrs (range 45–100 yrs) and the majority were put on comfort care with DNR orders in place. Among these patients 17 (70.8%) had DM, four (16.7%) were PreDM and three (12.5%) were nonDM.

### Covid-19 associated hyperglycemia

Obesity, PreDM and DM are typically associated with elevated blood glucose levels. We found blood glucose was increased in the majority of Covid-19 patients at admission with a mean of 179.9 mg/dL. For most patients, these values were obtained after fasting. The presenting FBGs were markedly elevated in many Covid-19 patients and 15 (10.2%) had FBG levels > 350 mg/dL on admission; four were in diabetic ketoacidosis.

While transient increases in blood glucose may be due to stress and more prolonged elevations can occur during treatment with corticosteroids, we found 23 of 54 (45.6%) Covid-19 patients with PreDM had persistently and markedly elevated FBG in the absence of corticosteroid therapy. Similarly, six patients without DM and with normal HbA1C levels also had repeatedly elevated FBG. Together, these 29 patients had FBG levels consistent with new onset DM and temporally associated with recent acquisition of SARS-CoV-2 infection. These findings support the possibility of direct dysregulation of glucose metabolism due to a newly acquired viral infection and point to an increased likelihood of developing severe Covid-19 in these high-risk patients.

## DISCUSSION

Identifying risk factors for development of severe Covid-19 in patients hospitalized with SARS-2-CoV is imperative for informing clinical decisions around patient care. Initial reports of Covid-19 from countries impacted early in the pandemic showed a strong association between older age and risk of Covid-19 mortality.(4,5) Data now emerging from the United States, which has the highest number of Covid-19 cases globally to date, has demonstrated increased incidence of severe Covid-19 in patients with co-morbid conditions including hypertension, obesity and diabetes. These findings have important implications for managing the clinical course of Covid-19 in severely ill patients and further understanding the pathophysiology of this disease.

Our single-center, consecutive series study of 184 patients with Covid-19 demonstrates several important findings. The majority of patients who developed moderate or severe Covid-19 had DM or preDM based on their HbA1C levels. Severe Covid-19, defined by the need for intubation, did not occur in the absence of DM, whether pre-existing or newly diagnosed, PreDM or obesity. Our data also establish that SARS-CoV-2 infection significantly worsens hyperglycemia in patients with glucose metabolism problems.

Our data in patients with severe Covid-19 and DM are consistent with a recent report by Bhatraju and colleagues.(8) In both studies, 58–62% of patients severely ill with Covid-19 were diabetic with mean BMIs > 30 and the majority had elevated blood glucose. Additionally, we found 24% of patients with moderate-severe Covid-19 in our study were prediabetic. Taken together these data suggest that insulin resistance and uncontrolled glycemia play a significant role in worsening Covid-19. In all critically ill Covid-19 patients, blood glucose levels were elevated and tight glycemic control may therefore be an important consideration for improving clinical outcomes.

Several studies on Covid-19 patients have reported on diabetes as a pre-existing diagnosis. In two recent observational studies, ∼36% of Covid-19 patients were diabetic.(16,17) These studies relied on passive surveillance at the time of admission. Similarly, 42.9% of our patients were known diabetics at the time of admission. However, we specifically reviewed prior medical records to ascertain each patient’s diabetes status. Further, we diagnosed an additional 17.1% during this admission. Most studies have not reported on prediabetes, a recognized syndrome with impaired glucose metabolism. By measuring HbA1C levels in every patient, we diagnosed 20.4% with prediabetes. Because of active surveillance, our data diabetes and prediabetes are replete and accurate.

The relationship of poor glycemic control and severe Covid-19 suggests a unique pathophysiological effect of SARS-CoV-2 infection on host metabolism. We have seen several non-diabetic Covid-19 patients with persistently high FBG levels. This is consistent with prior observations from the 2003 SARS-CoV outbreak, in which patients with pneumonia developed new onset DM and those with higher FBG levels had poor clinical outcomes.(9) SARS-CoV was also shown to cause new onset DM or glucose intolerance in patients.(18) The onset or worsening of hyperglycemia in patients with newly acquired SARS-CoV-2 infection suggests a similar mechanism may occur involving viral mediated disruption of glucose metabolism in Covid-19 patients.

It is unlikely the development of severe Covid-19 in these patients can be explained by the direct effect of changes in glucose metabolism on host immunity. To date, we have not seen Covid-19 patients with underlying diseases of immune dysregulation including AIDS or lupus. Nor have we seen Covid-19 patients with active lymphoma. Rather, these data suggest that SARS-CoV-2 infection is associated with physiologic changes in glucose metabolism that may permit the virus to replicate more efficiently.

Replication of SARS-CoV-2 in host cells is mediated by the ACE2 cell surface enzyme, which serves as the primary receptor for the virus.(19) As part of the renin-angiotensin system, ACE2 is expressed in many tissues, including pancreatic beta and apocrine cells. As noted in early reports from China, mild elevations of lipase produced by pancreatic apocrine cells may occur, and we have seen this in many, but not all, of our Covid-19 patients.(19) Clinically, SARS-CoV-2 appears to cause new or worsening hyperglycemia, which may lead to more severe pneumonia. In our experience, a tipping point is reached in Covid-19 patients who have symptoms lasting anywhere from two days to over three weeks and the disease then “takes off”. Hospitalization before this acceleration can reduce the rate of critical illness.

It is important to note that our study has several limitations. Patients were seen at a single clinical site and cared for by one group of clinicians. While it is possible our study population is disproportionately weighted towards patients with poor underlying health, the Covid-19 patients in this study were consecutive referrals to our service over the course of seven weeks in a suburban hospital. It is, therefore, unlikely that a selection bias exists, except for the criteria used by the admitting physicians. Diabetes itself was not considered a criterion for referral.

Given the urgency of finding solutions to this present crisis, our findings may assist in prognostication and triage decisions. Our data shed light on the impact of DM, preDM and uncontrolled hyperglycemia in driving severe Covid-19 and will facilitate identification of novel pathogenesis pathways associated with SARS-CoV-2 infection. This, in turn, may lead to new approaches to therapeutic intervention. Our data currently support the use of tight glycemic control in patients with hyperglycemia. Our observations are also in line with the WHO recommendation that corticosteroids not be used for COVID-19 pneumonia.

Finally, our findings caution that Covid-19 patients with DM, PreDM or obesity should be monitored closely. Those not infected should be particularly careful to avoid exposure to SARS-CoV-2. This information may be useful in healthcare and other settings to reduce the chances of infection in these high-risk individuals and, conversely, to help triage nonDM, normal glycemic Covid-19 patients safely and efficiently. [1–19]

## Data Availability

I can make the data available in spreadsheet format on Google.

